# Race and Socioeconomic Status Impact Survival from Early and Late-Onset Colorectal Cancer

**DOI:** 10.64898/2026.06.24.26356439

**Authors:** Kristen Purrington, Mei-Chin Hsieh, Siddhi Patil, Batsirai Mabvakure, Jaeil Ahn, Rui Zhang, Julie J. Ruterbusch, Rashmi Samdani, Grace Lee, Angela Wenzlaff, Sophia Latif, Chiranjeev Dash, Maureen Sartor, Ann G. Schwartz, Elena M. Stoffel, Laura S. Rozek

## Abstract

**Background:** Colorectal cancer (CRC) disproportionately affects non-Hispanic Black (NHB) Americans compared to Non-Hispanic White (NHW), with more cases arising before age 50. Racial disparities in outcomes reflect complex interactions among healthcare access, socioeconomic factors, and structural racism, yet analyses linking individual-level data for these factors to survival remain limited.

**Methods:** We examined overall and CRC-specific survival among NHB and NHW patients diagnosed between 2013 and 2022 enrolled in the Disparities and Cancer Epidemiology (DANCE) cohort, a population-based study of CRC in metropolitan Detroit and Louisiana. Multivariable Cox regression and competing-risks models were used to assess the roles of race, age of onset, neighborhood deprivation, and stage on survival outcomes.

**Results:** Among 1,019 CRC cases (57% NHB, 43% NHW), NHB patients were more likely to reside in high-deprivation neighborhoods, report lower household incomes, and present with right-sided tumors, though stage at diagnosis did not differ by race. In multivariable analysis, stage was the strongest predictor of survival, while neighborhood deprivation (per 10-unit ADI increase: HR = 1.14) was independently associated with worse survival; NHB race was not significantly associated with survival after adjustment. Younger age at diagnosis was associated with a survival advantage in regional-stage disease but paradoxically with worse survival in distant-stage disease, and higher deprivation predicted worse survival in both local and distant but not regional stage.

**Conclusion:** Our study shows that socioeconomic factors, as measured by ADI and household income, accounts for some, but not all, of the disparities in survival between NHB and NHW CRC cases.

## Background

Colorectal cancer (CRC) remains a significant health burden in the United States, ranking as the third most commonly diagnosed cancer and second leading cause of cancer-related death (1). Despite decades of progress driven by population-based screening and advances in treatment, the benefits of these improvements have not been distributed equitably. Racial disparities in CRC incidence, stage at diagnosis, and survival are well-documented and persistent (2), and an emerging shift in the age distribution of diagnoses threatens to further widen these gaps. Birth cohort analyses show a clear generational shift: people born since ∼1950–1960 in many countries face substantially higher CRC risk at a given age, with tumors arising in the distal colon and rectum at younger ages (3).

The changing epidemiology of CRC occurs in the context of long-standing racial and socioeconomic disparities. For non-Hispanic Black (NHB) Americans, who are 15% more likely to develop and 35% more likely to die from CRC than non-Hispanic White (NHW) Americans, the proportion of cases arising before age 50 (i.e., EOCRC: early-onset colorectal cancer) is already greater than in the general population (11% compared to 5%) (2). While EOCRC rates are converging due to a rapid rise among White patients, this does not mitigate the survival disadvantage that Black patients continue to experience, particularly considering that NHB patients are more likely to be diagnosed at a distant stage (4). CRC is the leading cause of cancer death in individuals under the age of 50 (5), and there is evidence from surveillance-based studies that NHB cases have worse survival from EOCRC (6–8).

Racial disparities in CRC outcomes reflect differences in access to financial and material resources, quality healthcare and screening, educational attainment, transportation, and the chronic physiologic burden of structural racism (9). For overall CRC, factors such as insurance coverage, treatment delays, and area-level deprivation account for a portion but do not fully explain racial disparities in CRC survival (7, 10, 11). For EOCRC specifically, the literature is more mixed with respect to the degree to which neighborhood-level socioeconomic factors explain racial differences (12, 13). Importantly, these registry-based studies were unable to account for key individual-level health behaviors and socioeconomic factors. Studies using individual-level and area-level SDOH data combined with robust clinical data are vital to taking the next step to identifying factors that influence outcomes in diverse populations. We conducted analyses on data collected as a part of the Disparities and Cancer Epidemiology (DANCE) study, a registry-based cohort study of colorectal cancer in metropolitan Detroit and Louisiana, to characterize survival in NHW and NHB CRC in the context of individual-level and area-level socioeconomic factors, clinical factors, and age.

## Methods

### Disparities and Cancer Epidemiology (DANCE) Cohort

The Disparities and Cancer Epidemiology cohort is a population-based colorectal cancer cohort designed to address the biologic basis of the demonstrated relationship between neighborhood deprivation and colorectal cancer. NHB and NHW CRC survivors were recruited through the Metropolitan Detroit Cancer Surveillance System (MDCSS) and the Louisiana Tumor Registry (LTR). A subset of DANCE cases are Detroit Research on Cancer Survivors (ROCS) cohort participants (n=456 NHB cases), described below. The catchment area of the DANCE study includes the Detroit metropolitan area in Michigan (Wayne, Oakland, and Macomb counties) and the entire state of Louisiana. Patients eligible for this study were diagnosed with CRC from 2013-2022. All DANCE participants completed baseline questionnaires either online, over the phone with a trained interviewer, or by returning a written copy. All DANCE participants provided informed consent, and study protocols were approved by Institutional Review Boards at the University of Michigan, Georgetown University, Wayne State University, and Louisiana State University.

### Research on Cancer Survivorship (ROCS) Cohort

Detroit ROCS is a population-based cohort designed to understand medical, behavioral, financial, and psychosocial outcomes among Black cancer survivors in metropolitan Detroit (14). Survivors diagnosed between ages 20 and 79 years were eligible to join the cohort if they were diagnosed with first primary invasive breast, colorectal, lung, or prostate cancer since January 1, 2013, and endometrial and other young onset (age at diagnosis ≥20 and ≤49) cancer cases diagnosed on or after January 1, 2016. Eligible participants were identified through the population-based MDCSS cancer registry and self-reported African American or Black race. Survivors diagnosed through July 2021 were eligible to join the ROCS cohort, and the last participants were enrolled in September 2022. At enrollment, participants completed a baseline survey online, over the phone, or using a mailed paper survey. The Institutional Review Board at Wayne State University approved this research.

### Clinical, demographic, health behavior, and area-level data

Clinical data were obtained through the MDCSS and LTR registries including age at diagnosis, sex, SEER tumor stage, primary site, first line radiation therapy, first line chemotherapy, vital status, length of active follow-up, and cause of death. Baseline questionnaires captured body mass index (BMI) at diagnosis, smoking status (defined as ever having smoked 100 cigarettes), insurance type, household income, alcohol use, and census tract at diagnosis. Area deprivation index (ADI) national percentile rankings (2015 version (created using 2011-2015 ACS data and updated to the v3.1 methodology); scale 0 (least disadvantaged) to 100 (most disadvantaged)) were obtained from The Neighborhood Atlas https://www.neighborhoodatlas.medicine.wisc.edu/ linked by census tract (15).

### Statistical Analyses

Demographic and clinical characteristics of CRC participants were summarized as means with standard deviation (SD) for continuous variables and counts with percentages for categorical variables. Group differences were assessed using Pearson’s chi-squared tests or exact binomial tests between NHB and NHW, low ADI (≤78 <=50) and high ADI (>78 >50), and among the two/three age at CRC onset groups (50 or over and <50 year, >65, 50 – 64 and <50 year), respectively. Overall survival (OS) was estimated and visualized using the Kaplan Meier method and log-rank tests were used to test differences by race, age, sex, ADI (binary), SEER stage at diagnosis, and primary sites. Multivariable Cox proportional hazard regression models were fitted adjusting for ADI (continuous), race, age, sex, BMI, income, SEER stage, and primary sites. To assess whether the association between ADI and OS was modified by race, SEER stage, or age of onset, Cox regression with two-way interactions were conducted. We further explored stratified Cox analyses classified by age onset, race, SEER stage, and ADI subgroups. Noting that a substantial proportion of deaths were non-CRC-specific, we additionally applied the nonparametric cumulative function of Fine and Gray (16) and fitted CRC cause-specific Cox proportional hazard models with identical covariate adjustments. The Schoenfeld residuals were evaluated to assess goodness-of-fits and the proportional hazard assumption. A two-sided p<0.05 was used to determine statistical significance. All analyses were conducted using SAS Version 9.4. (16)

## Results

Of the 1019 cases available, we categorized age into three groups representative of year of birth (Table 2): under age 50 (n = 180, 18%), 50 – 64 (n = 486, 48%), and 65 years of age and older (n = 353, 34%). Younger age categories were more likely to be obese (p = 0.004). The proportion of cases in the obese category was 32.9% in the >65 age group, 41.2% in the 50–64-year age group, and 51.1% in the under 50 age group. As age group decreased, the proportion of right-sided cancers decreased (54.7%, 35.6%, and 29.4% for the >65, 50 – 64, and <50 year age groups, respectively), the proportion of rectal cancers increased (22.1%, 27.8%, and 31.1% for the >65, 50 – 64, and <50 year age groups, respectively), and the proportion of cancers diagnosed at late stage increased (7.4%, 17.9%, and 23.9% for the >65, 50 – 64, and <50 year age groups, respectively).

We next considered race, 441 (43%) were NHW and 578 (57%) were NHB (Table 1). NHB cases were slightly more likely to be diagnosed between 50 and 64 in our cohort (50.7% vs 43.8%) and less likely to be diagnosed after 65 years (31.8% vs 31.3%, p = 0.061). There were several other notable differences by race in our cohort. NHB CRC cases were more likely to be female (p = 0.014), more likely to live in high deprivation areas (median ADI=95, p<0.001), less likely to have graduated from high school (p<0.001), and more likely to report annual household incomes less than $20,000 (p<0.001). Tumors from NHB CRC cases tended to be right-sided (46% vs 35%, p<0.001), although stage at diagnosis did not differ.

**Table 1.** Demographic and clinical characteristics of CRC patients by race.

| Variable |  | Total | NHW | NHB | P value |
| --- | --- | --- | --- | --- | --- |
| Total |  | 1019 | 441 (43.3%) | 578 (56.7%) |  |
| Age at diagnosis |  |  |  |  |  |
|  | Mean (STD) | 59.5 ± 10.7 | 60 ± 11.1 | 59.1 ± 10.3 | 0.15 |
|  | Median (Range) | 60 (23 - 79) | 61 (26 - 79) | 60 (23 - 79) |  |
| Age at diagnosis |  |  |  |  |  |
|  | 65+ | 353 (34.6%) | 169 (38.3%) | 184 (31.8%) | 0.061 |
|  | 50-64 | 486 (47.7%) | 193 (43.8%) | 293 (50.7%) |  |
|  | <50 | 180 (17.7%) | 79 (17.9%) | 101 (17.5%) |  |
| State |  |  |  |  |  |
|  | Michigan | 849 (83.3%) | 314 (71.2%) | 535 (92.6%) | <0.001 |
|  | Louisiana | 170 (16.7%) | 127 (28.8%) | 43 (7.4%) |  |
| Sex |  |  |  |  |  |
|  | Female | 491 (48.2%) | 193 (43.8%) | 298 (51.6%) | 0.014 |
|  | Male | 528 (51.8%) | 248 (56.2%) | 280 (48.4%) |  |
| National ADI |  |  |  |  |  |
|  | Mean (STD) | 69.6 ± 28.7 | 50.7 ± 26.1 | 84 ± 21.3 | <0.001 |
|  | Median (Range) | 78 (2 - 100) | 50 (2 - 100) | 95 (4 - 100) |  |
| National ADI |  |  |  |  |  |
|  | Low (≤78) | 511 (50.1%) | 355 (80.5%) | 156 (27.0%) | <0.001 |
|  | High (>78) | 508 (49.9%) | 86 (19.5%) | 422 (73.0%) |  |
| Body Mass Index at diagnosis |  |  |  |  |  |
|  | Normal (< 25) | 247 (24.2%) | 112 (25.4%) | 135 (23.4%) | 0.47 |
|  | Overweight (25 – < 30) | 333 (32.7%) | 148 (33.6%) | 185 (32.0%) |  |
|  | Obese (≥ 30) | 408 (40.0%) | 167 (37.9%) | 241 (41.7%) |  |
|  | Unknown | 31 (3.0%) | 14 (3.2%) | 17 (2.9%) |  |
| Highest Level Education |  |  |  |  |  |
|  | ≤High School/ GED | 327 (32.1%) | 110 (24.9%) | 217 (37.5%) | <0.001 |
|  | Some college(1-2yrs) | 374 (36.7%) | 151 (34.2%) | 223 (38.6%) |  |
|  | 4 yrs college/Graduate | 308 (30.2%) | 179 (40.6%) | 129 (22.3%) |  |
|  | Unknown | 10 (1.0%) | 1 (0.2%) | 9 (1.6%) |  |
| Last Year Household Income (before Taxes) |  |  |  |  |  |
| | < \$20000 | 292 (28.7%) | 49 (11.1%) | 243 (42.0%) | <0.001 |
| | \$20,000 - \$59,999 | 310 (30.4%) | 130 (29.5%) | 180 (31.1%) | |
| | ≥\$60,000 | 331 (32.5%) | 230 (52.2%) | 101 (17.5%) | |
|  | Unknown | 86 (8.4%) | 32 (7.3%) | 54 (9.3%) |  |
| Smoked 100 Cigarettes in lifetime |  |  |  |  |  |
|  | No | 541 (53.1%) | 230 (52.2%) | 311 (53.8%) | 0.87 |
|  | Yes | 460 (45.1%) | 198 (44.9%) | 262 (45.3%) |  |
|  | Unknown | 18 (1.8%) | 13 (2.9%) | 5 (0.9%) |  |
| Alcohol User in last 4 weeks |  |  |  |  |  |
|  | No | 552 (54.2%) | 208 (47.2%) | 344 (59.5%) | <0.001 |
|  | Yes | 452 (44.4%) | 225 (51.0%) | 227 (39.3%) |  |
|  | Unknown | 15 (1.5%) | 8 (1.8%) | 7 (1.2%) |  |
| Primary Site |  |  |  |  |  |
|  | Left Colon | 302 (29.6%) | 145 (32.9%) | 157 (27.2%) | 0.001 |
|  | Rectum & Rectosigmoid J. | 269 (26.4%) | 133 (30.2%) | 136 (23.5%) |  |
|  | Right Colon | 419 (41.1%) | 153 (34.7%) | 266 (46.0%) |  |
|  | Other | 29 (2.8%) | 10 (2.3%) | 19 (3.3%) |  |
| SEER Stage at Dx |  |  |  |  |  |
|  | Local | 360 (35.3%) | 145 (32.9%) | 215 (37.2%) | 0.15 |
|  | Regional | 503 (49.4%) | 233 (52.8%) | 270 (46.7%) |  |
|  | Distant | 156 (15.3%) | 63 (14.3%) | 93 (16.1%) |  |
| Radiation as 1st course of Rx |  |  |  |  |  |
|  | No/Unknown | 951 (93.3%) | 432 (98.0%) | 519 (89.8%) | <0.001 |
|  | Yes | 68 (6.7%) | 9 (2.0%) | 59 (10.2%) |  |
| Chemotherapy as 1st course Rx |  |  |  |  |  |
|  | No/Unknown | 734 (72.0%) | 408 (92.5%) | 326 (56.4%) | <0.001 |
|  | Yes | 285 (28.0%) | 33 (7.5%) | 252 (43.6%) |  |
| Vital Status |  |  |  |  |  |
|  | Deceased | 226 (22.2%) | 61 (13.8%) | 165 (28.5%) | <0.001 |
|  | Alive | 793 (77.8%) | 380 (86.2%) | 413 (71.5%) |  |

**Table 2.** Demographic and clinical characteristics of CRC patients by age.

| Variable | Total | 65 + | 50 - 64 | < 50 | P value |
| --- | --- | --- | --- | --- | --- |
| Total | 1019 | 353 (34.6%) | 486 (47.7%) | 180 (17.7%) |  |
| Age at diagnosis |  |  |  |  |  |
| Mean (STD) | 59.5 ± 10.7 | 70.7 ± 4.05 | 57.5 ± 4.36 | 42.9 ± 5.43 | <0.001 |
| Median (Range) | 60 (23 - 79) | 70 (65 - 79) | 58 (50 - 64) | 44 (23 - 49) |  |
| Race |  |  |  |  |  |
| NHW | 441 (43.3%) | 169 (47.9%) | 193 (39.7%) | 79 (43.9%) | 0.061 |
| NHB | 578 (56.7%) | 184 (52.1%) | 293 (60.3%) | 101 (56.1%) |  |
| State |  |  |  |  |  |
| Michigan | 849 (83.3%) | 283 (80.2%) | 410 (84.4%) | 156 (86.7%) | 0.11 |
| Louisiana | 170 (16.7%) | 70 (19.8%) | 76 (15.6%) | 24 (13.3%) |  |
| Sex |  |  |  |  |  |
| Female | 491 (48.2%) | 179 (50.7%) | 225 (46.3%) | 87 (48.3%) | 0.45 |
| Male | 528 (51.8%) | 174 (49.3%) | 261 (53.7%) | 93 (51.7%) |  |
| National ADI |  |  |  |  |  |
| Mean (STD) | 69.6 ± 28.7 | 69.6 ± 27.7 | 70.3 ± 29.7 | 67.6 ± 27.7 | 0.55 |
| Median (Range) | 78 (2 - 100) | 78 (2 - 100) | 80.5 (2 - 100) | 72 (4 - 100) |  |
| National ADI |  |  |  |  |  |
| Low (≤78) | 511 (50.1%) | 177 (50.1%) | 234 (48.1%) | 100 (55.6%) | 0.24 |
| High (>78) | 508 (49.9%) | 176 (49.9%) | 252 (51.9%) | 80 (44.4%) |  |
| Body Mass Index |  |  |  |  |  |
| Normal (< 25) | 247 (24.2%) | 93 (26.3%) | 121 (24.9%) | 33 (18.3%) | 0.004 |
| Overweight (25 – < 30) | 333 (32.7%) | 127 (36.0%) | 154 (31.7%) | 52 (28.9%) |  |
| Obese (≥ 30) | 408 (40.0%) | 116 (32.9%) | 200 (41.2%) | 92 (51.1%) |  |
| Unknown | 31 (3.0%) | 17 (4.8%) | 11 (2.3%) | 3 (1.7%) |  |
| Highest Level Education |  |  |  |  |  |
| ≤High School/ GED | 327 (32.1%) | 120 (34.0%) | 161 (33.1%) | 46 (25.6%) | 0.31 |
| Some college(1-2yrs) | 374 (36.7%) | 130 (36.8%) | 172 (35.4%) | 72 (40.0%) |  |
| 4 yrs college/Graduate | 308 (30.2%) | 101 (28.6%) | 146 (30.0%) | 61 (33.9%) |  |
| Unknown | 10 (1.0%) | 2 (0.6%) | 7 (1.4%) | 1 (0.6%) |  |
| Last Year Household Income<br>(before Taxes) |  |  |  |  |  |
| < \$20000 | 292 (28.7%) | 87 (24.6%) | 160 (32.9%) | 45 (25.0%) | <0.001 |
| \$20,000 - \$59,999 | 310 (30.4%) | 134 (38.0%) | 129 (26.5%) | 47 (26.1%) | |
| ≥\$60,000 | 331 (32.5%) | 91 (25.8%) | 165 (34.0%) | 75 (41.7%) | |
| Unknown | 86 (8.4%) | 41 (11.6%) | 32 (6.6%) | 13 (7.2%) |  |
| Smoked 100 Cigarettes in lifetime |  |  |  |  |  |
| No | 541 (53.1%) | 168 (47.6%) | 256 (52.7%) | 117 (65.0%) | <0.001 |
| Yes | 460 (45.1%) | 178 (50.4%) | 223 (45.9%) | 59 (32.8%) |  |
| Unknown | 18 (1.8%) | 7 (2.0%) | 7 (1.4%) | 4 (2.2%) |  |
| Alcohol User in last 4 weeks |  |  |  |  |  |
| No | 552 (54.2%) | 221 (62.6%) | 247 (50.8%) | 84 (46.7%) | <0.001 |
| Yes | 452 (44.4%) | 127 (36.0%) | 233 (47.9%) | 92 (51.1%) |  |
| Unknown | 15 (1.5%) | 5 (1.4%) | 6 (1.2%) | 4 (2.2%) |  |
| Primary Site |  |  |  |  |  |
| Left Colon | 302 (29.6%) | 77 (21.8%) | 159 (32.7%) | 66 (36.7%) | <0.001 |
| Rectum & Rectosigmoid J. | 269 (26.4%) | 78 (22.1%) | 135 (27.8%) | 56 (31.1%) |  |
| Right Colon | 419 (41.1%) | 193 (54.7%) | 173 (35.6%) | 53 (29.4%) |  |
| Other | 29 (2.8%) | 5 (1.4%) | 19 (3.9%) | 5 (2.8%) |  |
| SEER Stage at Dx |  |  |  |  |  |
| Local | 360 (35.3%) | 144 (40.8%) | 169 (34.8%) | 47 (26.1%) | <0.001 |
| Regional | 503 (49.4%) | 183 (51.8%) | 230 (47.3%) | 90 (50.0%) |  |
| Distant | 156 (15.3%) | 26 (7.4%) | 87 (17.9%) | 43 (23.9%) |  |
| Radiation as 1st course of Rx |  |  |  |  |  |
| No/Unknown | 951 (93.3%) | 331 (93.8%) | 451 (92.8%) | 169 (93.9%) | 0.81 |
| Yes | 68 (6.7%) | 22 (6.2%) | 35 (7.2%) | 11 (6.1%) |  |
| Chemotherapy as 1st course Rx |  |  |  |  |  |
| No/Unknown | 734 (72.0%) | 281 (79.6%) | 341 (70.2%) | 112 (62.2%) | <0.001 |
| Yes | 285 (28.0%) | 72 (20.4%) | 145 (29.8%) | 68 (37.8%) |  |
| Vital Status |  |  |  |  |  |
| Deceased | 226 (22.2%) | 76 (21.5%) | 109 (22.4%) | 41 (22.8%) | 0.93 |
| Alive | 793 (77.8%) | 277 (78.5%) | 377 (77.6%) | 139 (77.2%) |  |

While overall survival did not differ by age group (log rank p = 0.83), NHB CRC cases had worse overall survival than NHW in our cohort (log rank p value = <0.001) (Table 1). In univariate survival analyses, higher ADI ((≤78 <u><=50</u> vs (>78 <u>>50</u>), (p<0.001), distant and regional stage (p<0.001), NHB race (p<0.001), and male gender (p = 0.039) were associated with worse CRC overall survival. Survival did not differ by site of diagnosis in our cohort. We then considered multivariable adjusted models that characterized age (in three birth cohorts), race, and ADI as main effects (Figure 1). The largest predictor of overall survival was stage, with distant (HR = 11.61, 95% CI 7.73 – 17.4) and regional (Hazard ratio (HR) = 2.16, 95% CI 1.43 – 3.19) stages having significantly higher mortality than local stage disease. ADI was also significantly associated with higher mortality (HR = 1.14, 95% CI 1.06 – 1.22 for a 10-unit increase in ADI), while high income (>$60,000 annual income) was weakly associated with a survival advantage (HR = 0.66, 95% CI 0.43 – 1.02). Male CRC cases experienced worse survival (HR = 1.40, 95% CI 1.06 – 1.86). Interestingly, NHB race was not associated with survival in the multivariable model (HR = 1.10, 95% CI 0.75 – 1.61). Subdistribution hazard ratios from a Fine-Gray competing risk model show a reduced and nonsignificant association between male sex and survival from colorectal cancer (1.19 (0.83 -1.72).

**Figure 1.**
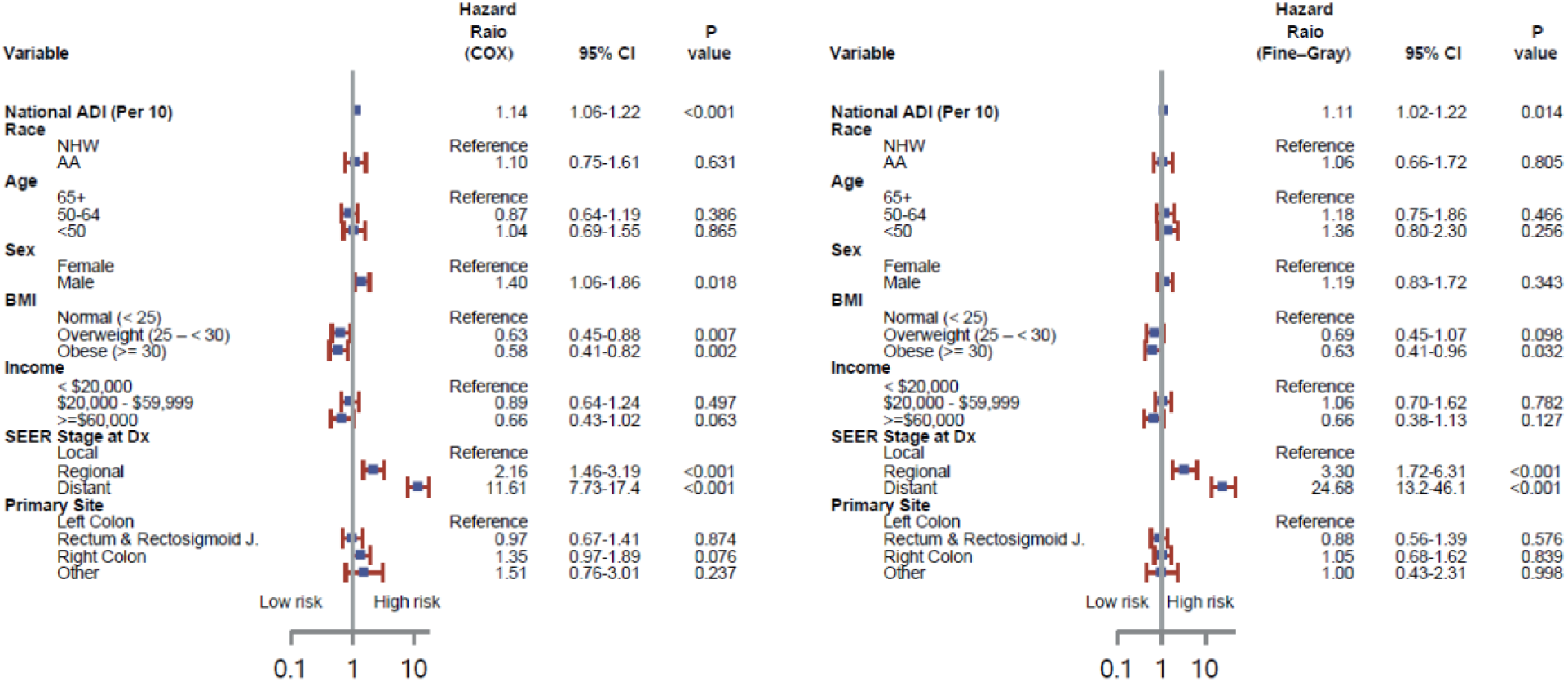
Multivariable-adjusted hazard ratios for overall survival in colorectal cancer patients. Hazard ratios (HRs) and 95% confidence intervals (CIs) are shown from Cox proportional hazards models and Fine-Gray competing risk models. The vertical line indicates HR = 1.

We stratified by stage to better understand within-group predictors of survival (Figure 2). Compared to cases diagnosed after age 65, younger age was associated with a protective effect for regional disease (50 – 64 years: HR = 0.53, 95% CI 0.34 – 0.84; <50 years: HR = 0.48, 95% CI 0.24 – 0.96). However, younger age was associated with a higher hazard of death when diagnosed at distant stage compared to cases older than 65 years diagnosed with distant disease (50-64 years: HR = 2.09, 95% CI 1.04 – 4.22; <50 years: HR = 2.43, 95% CI 1.12 – 5.27). A ten unit (%) increase in ADI was significantly associated with overall survival among local stage (HR = 1.37, 95% CI 1.09 – 1.73) and distant stage (1.17, 95% CI 1.04 – 1.31) but not regional stage disease (HR = 1.05, 95% CI 0.95 – 1.17). Household income was only associated with overall survival among regional stage disease, with a borderline protective effect in the $20,000 to $59,999 group (HR = 0.62, 95% CI 0.37 – 1.03 1.33) and a statistically significant effect in the ≥ $60,000 group (HR = 0.29, 95% CI 0.13 – 0.61). NHB race was not associated with a significant hazard of death in any stage group but was suggestive in distant stage (HR = 1.43, 95% CI 0.75 – 2.73). Male CRC cases had a higher hazard of death in the local stage compared (HR = 2.37, 95% CI 1.11 – 5.07) and regional stage groups (HR = 1.82, 95% CI 1.19 – 2.79) but not the distant stage group (HR = 1.05, 95% CI 0.66 – 1.67). In post-hoc analyses, we combined local and regional stages to estimate Fine-Gray competing risk models for early-stage cancer due to small numbers of deaths due to cancer. While age group was protective in overall models, there was no association with cause-specific survival. The estimates for distant stage did not differ substantially in competing risk models.

**Figure 2.**
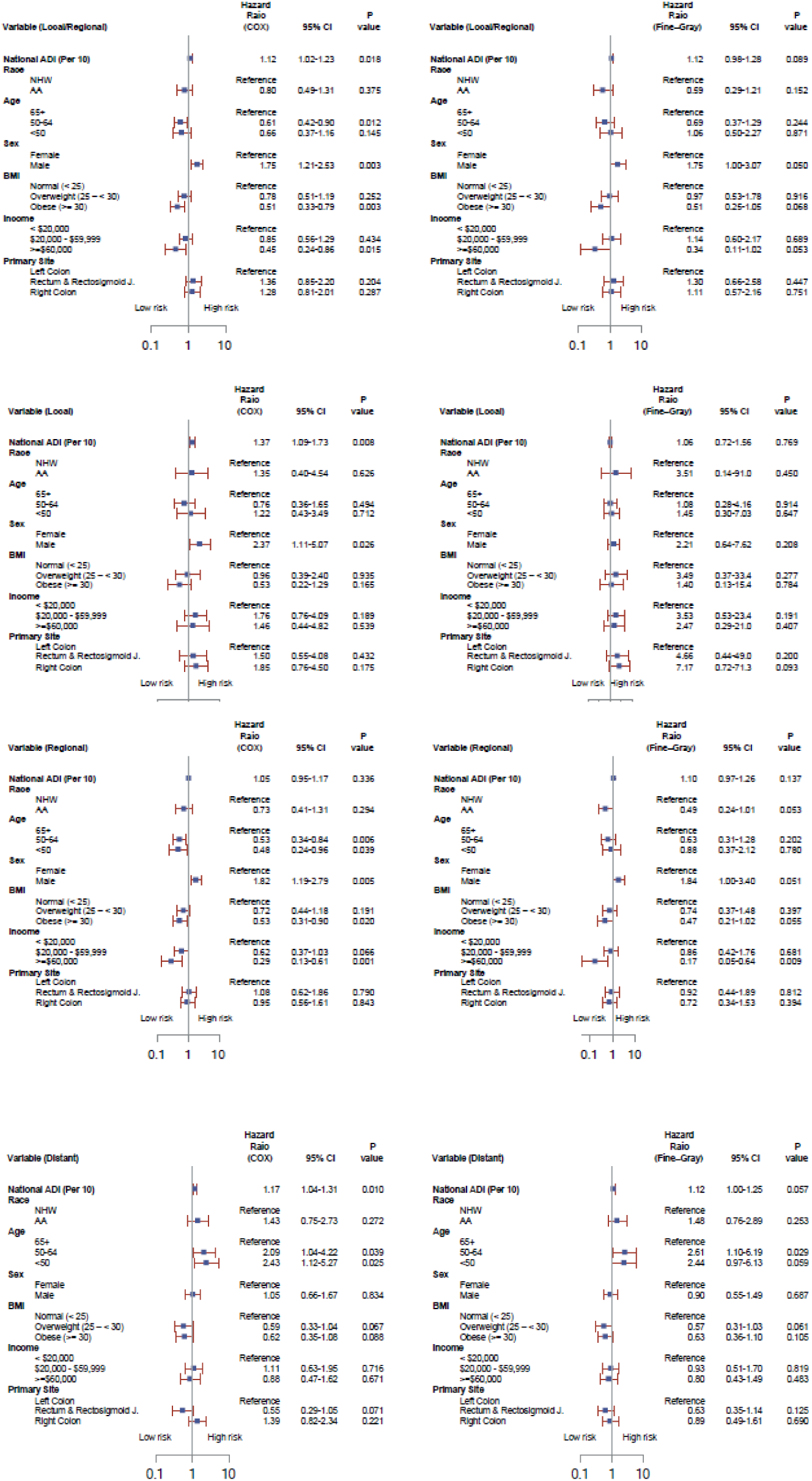
Multivariable-adjusted hazard ratios for overall survival in colorectal cancer patients by SEER stage group. Hazard ratios (HRs) and 95% confidence intervals (CIs) are shown from Cox and Fine-Gray models stratified by local, regional, distant, and combined local/regional stage groups. The vertical line indicates HR = 1.

Given the strong association between ADI and survival, we wanted to explore within-group variation with respect to ADI. When stratified by median ADI (ADI ≤78 and > 78, Figure 3), NHB race was borderline associated with an increased hazard of death in the ≤78 ADI group (HR = 1.64, 95% CI 0.97 – 2.78), but not the <u>≥</u>78 ADI group (HR = 1.15, 95% CI 0.68 – 1.95). Regional and distant stage were associated with an increased hazard in the ADI ≤78 group (Regional vs. local: HR = 3.61, 95% CI 1.66 – 7.84; Distant vs local: HR = 17.63, 95% CI 7.71 – 40.3) and the ADI <u>≥</u>78 group (Regional vs. local: HR = 1.73, 95% CI 1.09 – 2.75; Distant vs local: HR = 9.75, 95% CI 6.06 – 15.7). BMI had a protective effect in the ADI ≤78 and ADI <u>≥</u>78 for both overweight (HR=0.55, 95% CI 0.31-0.97) and obese (HR=0.46, 95% CI 0.24-0.85) groups compared to normal weight. Subdistribution hazard ratios from a Fine-Gray competing risk model are similar in both ADI strata, except the relationship with obesity is less profound in the <u>≥</u>78 ADI group.

**Figure 3.**
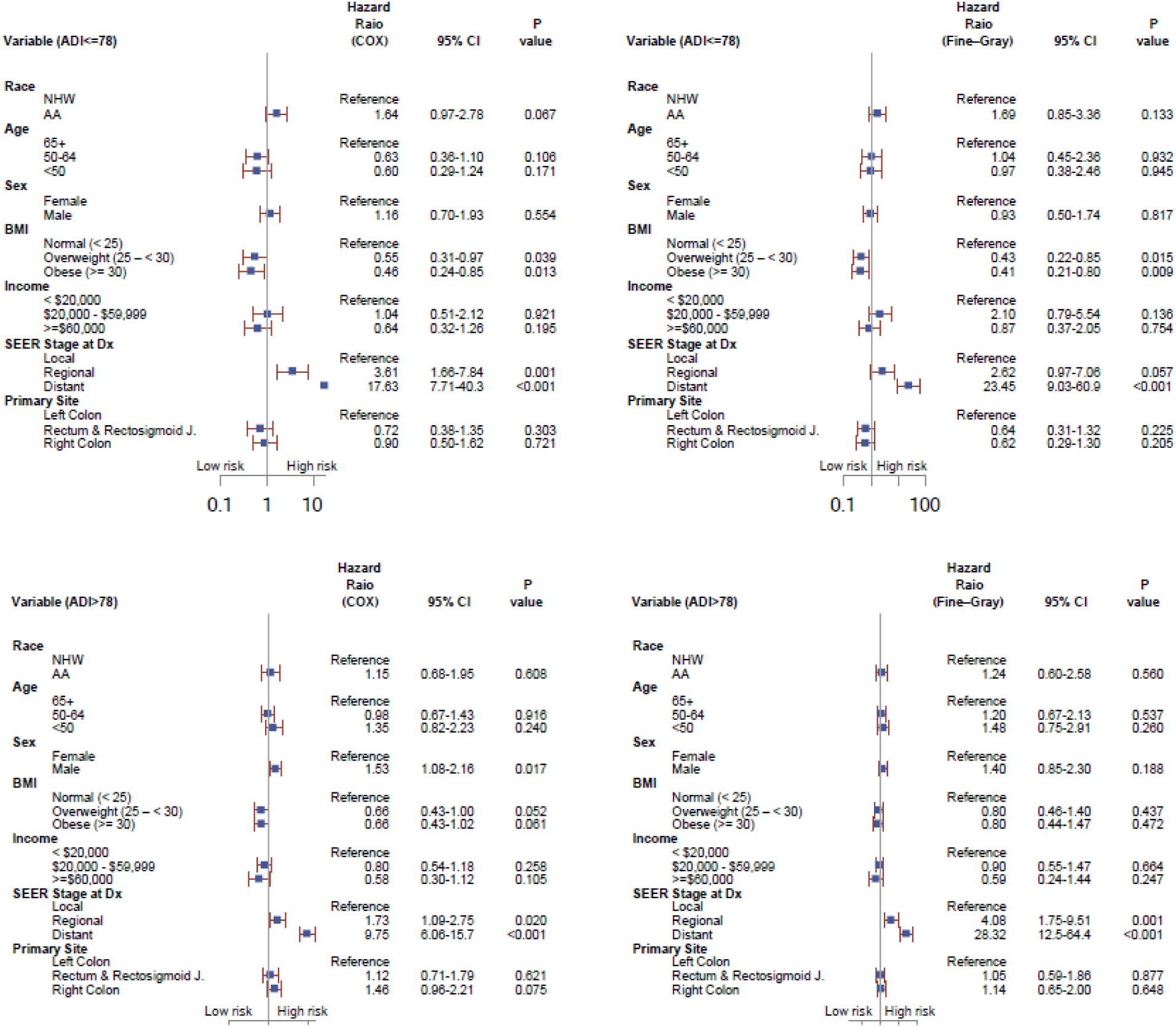
Multivariable-adjusted hazard ratios for overall survival in colorectal cancer patients stratified by area deprivation index (ADI). Hazard ratios (HRs) and 95% confidence intervals (CIs) are shown from Cox and Fine-Gray models for ADI ≤78 and ADI >78. The vertical line indicates HR = 1.

## Discussion

Even after adjustment for ADI and race, in the distant stage, younger age groups showed worse survival from CRC, potentially indicating a more aggressive disease in earlier age cohorts.

Age-period-cohort analyses in the US indicate that birth cohorts as early as the 1940-1950 cohort experienced an increase in EOCRC, indicating the likely influence of lifestyle and environmental changes on EOCRC over time (17). As such, we stratified our cohort into three age groups (<50 years, 50 – 64, and 65 and older) to reflect the changing epidemiology of CRC over time. In our cohort, stage at diagnosis increased in decreasing age groups, which is expected given potential screening rates in younger populations despite guidelines. We also note increasing proportion of left-sided and rectal tumors in younger age cohorts, with the 50 – 64-year age group with intermediate proportion of left-sided and rectal cancers to the oldest and younger cohorts. The proportion of cases in the obese category increased with younger birth cohorts and the proportion of cases reporting smoking decreased and alcohol use increased. All of these could be a reflection of cohort-specific differences but nevertheless may influence risk of and survival from CRC. Indeed, we found that overall survival differences were not different, but when stratified by stage, younger age was protective in CRCs diagnosed at a regional stage but associated with a higher hazard of death at distant stage. Based on our findings, we recommend that analyses consider age cohorts, not a binary age cutoff at 50, in analyzing CRC data.

Overall, our study also shows that socioeconomic status, as measured by ADI and household income, accounts for some, but not all, of the disparity in survival between NHB and NHW CRC cases. While univariate survival analyses indicated differences between NHB and NHW CRC cases, models that account for ADI and stage – both of which were associated with survival in univariate analyses – NHB race was no longer significantly associated with survival from CRC. In multivariable adjusted models, stage was the strongest predictor of survival, and ADI remained significant as well as higher household income, indicating that both area-level and individual-level indicators of socioeconomic status predicted overall survival. Interestingly, male gender was associated with worse survival in most models. We ran models by stage and ADI category to look at within-group predictors of survival and found variation that was not apparent in overall cohort analyses. Area-level SES (ADI) was significantly associated with survival in patients diagnosed with local and distant disease, but individual-level SES (household income) was associated with regional disease. Age group was protective for regional disease but was associated with a survival disadvantage in patients diagnosed with distant disease. NHB race was not significantly associated with survival in any stratified models, but NHB race was marginally associated with an increased hazard of death in the low ADI group.

Our study adds to the body of literature seeking to understand racial disparities in CRC outcomes. As there are few cohort studies that have an appreciable number of NHB subjects, most of the evidence is derived from SEER data and other large sources. These studies show worse survival in NHB cases and other minoritized populations in the US (8, 13, 18, 19). SEER-based and other registry studies offer the advantage of large numbers and harmonized data collection. They have also been informative of the geographic differences in CRC mortality across the United States. CRC mortality is highest in the southern US, with county-level hotspots in CRC mortality in the southeast and eastern Midwest (20, 21). EOCRC incidence is the highest in the south but rising in the western part of the US (22). Given the variation in CRC mortality rates, social determinants, and racial distribution across the US and within individual states, observational studies are a critical next step in characterizing the complex relationship between SES, race, and CRC survival.

Our study shows that accounting for SES – both area-level as defined by ADI and individual-level captured by household income – accounts for some but not all of the racial disparities in CRC survival in our population. However, we found that NHB CRC patients living in areas of less deprivation experienced worse survival than NHW patients, even when accounting for household income, suggesting that socioeconomic disadvantage alone does not capture the cumulative effects of systemic racism on healthcare access, quality of care, and chronic stress-related physiological burden. These findings are consistent with the diminishing returns hypothesis, where minoritized people do not experience the cancer survival benefits appreciated by non-Hispanic whites that has been characterized for several cancers in the US (23). Our data are limited by the narrow range of ADI scores overall and even more so among NHB CRC cases, supporting the need for more extensive studies that can evaluate the full range of ADI and individual factors across populations. Aggressive tumor phenotypes could also partially explain findings such as these, and our results support further, more intensive study of the tumor biology of CRC among NHB patients. While NHB CRC is underrepresented in the TCGA, previous somatic mutational profile analyses suggest the potential for a more aggressive phenotype in NHB patients (24–26).

Our results also indicate that males consistently have worse survival even when accounting for age, SES, and race. There is significant heterogeneity in cancer risk factors and survival between males and females, including a preponderance of left sided cancers and a smaller proportion of MSI high cancers in males (27). Similarly, a large SEER-based U.S. study of EOCRC found men had significantly worse overall, cancer-specific, and non-cancer survival than women, and male sex remained an independent adverse prognostic factor after multivariable adjustment and propensity matching (28). Higher number of comorbidities, individual lifestyle behaviors, and biologic differences have been hypothesized to contribute to observed gender survival differences and studies should address these differences in order to design effective interventions to improve survival in men (27).

This study has several strengths and limitations. Ascertainment was carried out in two diverse areas of the country which is reflected in the makeup of our cohort. Louisiana and metropolitan Detroit represent areas of our country with a high proportion of both African Americans and high disparities. As such, we are able to perform subgroup analyses that include adequate numbers of both NHB and NHW participants. Because subjects were ascertained using population-based registries, our sampling frame is unbiased. Race and socioeconomic status have a complex relationship with respect to survival from CRC. While our cohort has a high number of African American CRC cases, there are notable differences that make interpreting the data difficult. While approximately half of our NHW cases are in the high ADI category, 89% of our NHB cases are in that category. We also do not have an appreciable number of cases from Louisiana that would permit a subgroup analysis. Our ascertainment strategies are based on the successful Detroit Research on Cancer Survivorship (detroitrocs.org) study.

In conclusion, our data show that SES, at both area- and individual-levels, accounts for a substantial proportion of observed racial differences in CRC survival, although stratified analyses show the complexity of the relationship. Our data also support the idea that age group at diagnosis may correspond to generational changes in lifestyle and environmental contributions to CRC risk and survival during the 20^th^ century. Ongoing studies of CRC, race, and age will benefit from molecular characterization to define the biologic contribution to cancers in these populations.

## Acknowledgements

The funder did not play a role in the design of the study; the collection, analysis, and interpretation of the data; the writing of the manuscript; and the decision to submit the manuscript for publication.

## Data Availability Statement

The data underlying this study have already been collected and are not publicly available due to privacy and institutional restrictions. Access to the data can be requested with a justified research purpose by contacting the primary investigators, Dr. Laura Rozek, subject to institutional approval.

## Informed consent

Written informed consents were obtained from the human participants, and the study was conducted in accordance with the U.S. Common Rule. Additionally, the study was approved by the University of Michigan’s single Institutional Review Board (IRB) and by the IRBs at Georgetown University, Wayne State University, and Louisiana State University.

IRB: University of Michigan’s single IRB number: HUM00196924

